# Genetic and environmental etiology of the broad avoidant restrictive food intake disorder phenotype in 6- to-12-year-old Swedish twins

**DOI:** 10.1101/2022.09.08.22279706

**Authors:** Lisa Dinkler, Paul Lichtenstein, Sebastian Lundström, Henrik Larsson, Nadia Micali, Mark J. Taylor, Cynthia M. Bulik

## Abstract

**IMPORTANCE:** Avoidant restrictive food intake disorder (ARFID) is characterized by an extremely limited range and/or amount of food eaten, resulting in the persistent failure to meet nutritional and/or energy needs. Its etiology is poorly understood and knowledge of genetic and environmental contributions to ARFID is needed to guide future research.

**OBJECTIVE:** To determine the extent to which genetic and environmental factors contribute to the liability to the broad ARFID phenotype.

**DESIGN, SETTING, AND PARTICIPANTS:** A nationwide Swedish twin cohort including 16,951 twin pairs born 1992-2010 whose parents participated in the Child and Adolescent Twin Study in Sweden (CATSS) at twin age 9 or 12 years (49.4% female). CATSS was linked to the National Patient Register (NPR) and the Prescribed Drug Register (PDR).

**MAIN OUTCOME/MEASURES:** From CATSS, NPR, and PDR, we extracted all parent-reports, diagnoses, procedures, and prescribed drugs between age 6 and 12 that were relevant to the DSM-5 ARFID criteria and developed a composite measure for the ARFID phenotype (i.e., avoidant/restrictive eating with clinically significant impact such as low weight or nutritional deficiency, and with fear of weight gain as an exclusion). In sensitivity analyses, we controlled for autism and medical conditions that could account for the eating disturbance. We fitted univariate liability threshold models to estimate the relative contribution of genetic and environmental variation to the liability to the ARFID phenotype.

**RESULTS:** We identified 667 children (2.0%, 38.2% female) with the ARFID phenotype between age 6 and 12. Variation in the liability to ARFID was largely explained by additive genetic factors (0.79, 95% confidence interval [CI] 0.71-0.86), with significant contributions from non-shared environmental factors (0.21, 95% CI 0.14-0.29). Heritability was very similar when excluding children with autism (0.77, 95% CI 0.67-0.84) or medical illnesses that could account for the eating disturbance (0.80, 95% CI 0.71-0.86).

**CONCLUSIONS AND RELEVANCE:** Prevalence and sex distribution of the broad ARFID phenotype were similar to previous studies, supporting the use of existing epidemiological data to identify ARFID. This first study of the genetic and environmental etiology of ARFID suggests that ARFID is highly heritable, encouraging future twin and molecular genetic studies.

## Introduction

ARFID is a serious feeding and eating disorder formally recognized in DSM-5^1^ in 2013, and first included the ICD in 2022 (ICD-11).^2^ Characterized by an extremely limited *range* or *amount* of food consumed, and resulting in persistent failure to meet nutritional and/or energy needs, ARFID is associated with considerable individual, family, and social impairment,^3^ and medical consequences^4,5^ can be life-threatening. Unlike anorexia nervosa, dietary restriction in ARFID is *not* motivated by body image concerns or drive for thinness, but rather based on *sensory sensitivity to food qualities* (e.g., texture, smell, taste), *lack of interest in food/eating* (i.e., low appetite), and/or *fear of aversive somatic consequences of food intake* (e.g., choking, vomiting, allergic reactions),^1^ often in response to aversive eating experiences.^6^ With an estimated prevalence of 1-5%,^7,8^ ARFID is at least as common as autism^9^ and potentially as common as attention deficit hyperactivity disorder.^10^

The etiology of ARFID remains poorly understood and the genetics of ARFID remain understudied. Other eating disorders such as anorexia nervosa and bulimia nervosa have been shown to have moderate to high heritability,^11^ and large-scale genome-wide association studies (GWAS) have successfully identified risk loci for anorexia nervosa, and underscored the importance of considering metabolic factors in its etiology.^12^ In contrast, the heritability of ARFID is as yet unknown, although twin studies have been conducted on related phenotypes showing low to moderate heritabilities for macronutrient, micronutrient, and overall caloric intake (.21-.48)^13^ and fruit/vegetable liking (.37-.54);^14,15^ and moderate to high heritabilities for food fussiness (.46-.78),^14,16^ food neophobia (.58-.78),^16-18^ and appetite (.53-.84^19^; for a comprehensive review see^20^).

Importantly, the genetic epidemiology of ARFID is unknown because validated ARFID screening instruments are only starting to emerge. Until such measures have been developed and deployed, we can optimize available resources such as those held by the Swedish Twin Registry to create a diagnostic algorithm to identify an ARFID phenotype and study its prevalence, correlates, and etiology. The **aim of this study** was to determine the extent to which genetic and environmental factors contribute to the liability to ARFID. Based on the moderate to high heritability of other eating disorders (anorexia nervosa .48-.74, bulimia nervosa .55-.61, and binge-eating disorder .39-.45)^21^ and the above reported heritability estimates of ARFID-related traits, we expected at least moderate heritability of ARFID.

## Methods

### Participants

We leveraged existing data from the Child and Adolescent Twin Study in Sweden (CATSS), targeting all twins born in Sweden since July 1, 1992.^22^ CATSS is one of the largest twin studies in the world, contains a broad range of psychiatric and neurodevelopmental phenotypes, and is linked to the national population health and quality registers.^22^ Parents of twins are first invited to participate in CATSS at twin age 9 (the cohorts born July 1992 to June 1995 were assessed at age 12, CATSS-9/12). Zygosity was ascertained either using a panel of 48 single nucleotide polymorphisms, and/or an algorithm of five questions regarding twin similarity. CATSS and its linkage to the Swedish health registries was approved by the Regional Ethical Review Board in Stockholm, Sweden (02-289, 03-672, 2010/322-31/2, 2010/597-31/1, 2016/2135-31, 2018/1398-32).

This study included twins born 1992-2010 who were part of CATSS-9/12 (response rate ∼69%). For this sample, data from the National Patient Register (NPR; diagnostic and procedure codes from inpatient care with full coverage since 1987 and ∼80% of specialized outpatient care since 2001;^23^ ICD-9 codes 1987–1996, ICD-10 codes since 1997) was available until the end of 2016 and data from the Prescribed Drug Register (PDR; all dispensations of prescribed drugs since 2005, active drug ingredients coded according to the Anatomical Therapeutic Chemical [ATC] Classification System) was available until the end of 2017.^24^ We excluded twins with unknown zygosity (n=435) and missing co-twin (n=45). The final sample comprised 32,902 individuals (49.4% female; 81.1% age 9 and 18.9% age 12; i.e., 16,951 twin pairs (5,184 monozygotic [MZ], 5,936 dizygotic same-sex [DZ-ss], and 5,831 dizygotic opposite-sex [DZ-os]).

### Identification of the ARFID phenotype

To identify children with an ARFID phenotype, we extracted all information relevant to the DSM-5 criteria for ARFID from CATSS, NPR, and PDR, and developed a composite measure including indicators of avoidant/restrictive eating and potential clinically significant consequences of the eating behavior (e.g., low weight/failure to thrive, nutritional deficiency, nutritional supplements, psychosocial impairment; *DSM-5 ARFID criterion A*), while considering exclusion criteria such as body image concerns (e.g., fear of weight gain) and medical conditions that could potentially explain the eating disturbance (*DSM-5 ARFID criteria C & D*; **Figure 1**). *DSM-5 ARFID criterion B* (eating disturbance is not better explained by lack of available food or an associated culturally sanctioned practice) could not be considered as such information was not available. Furthermore, the clinical diagnoses of feeding and eating disorders and the specific CATSS items used to identify the ARFID phenotype are unlikely to reflect cultural practices causing the eating disturbance (**eTable 1**).

**Figure 1.**
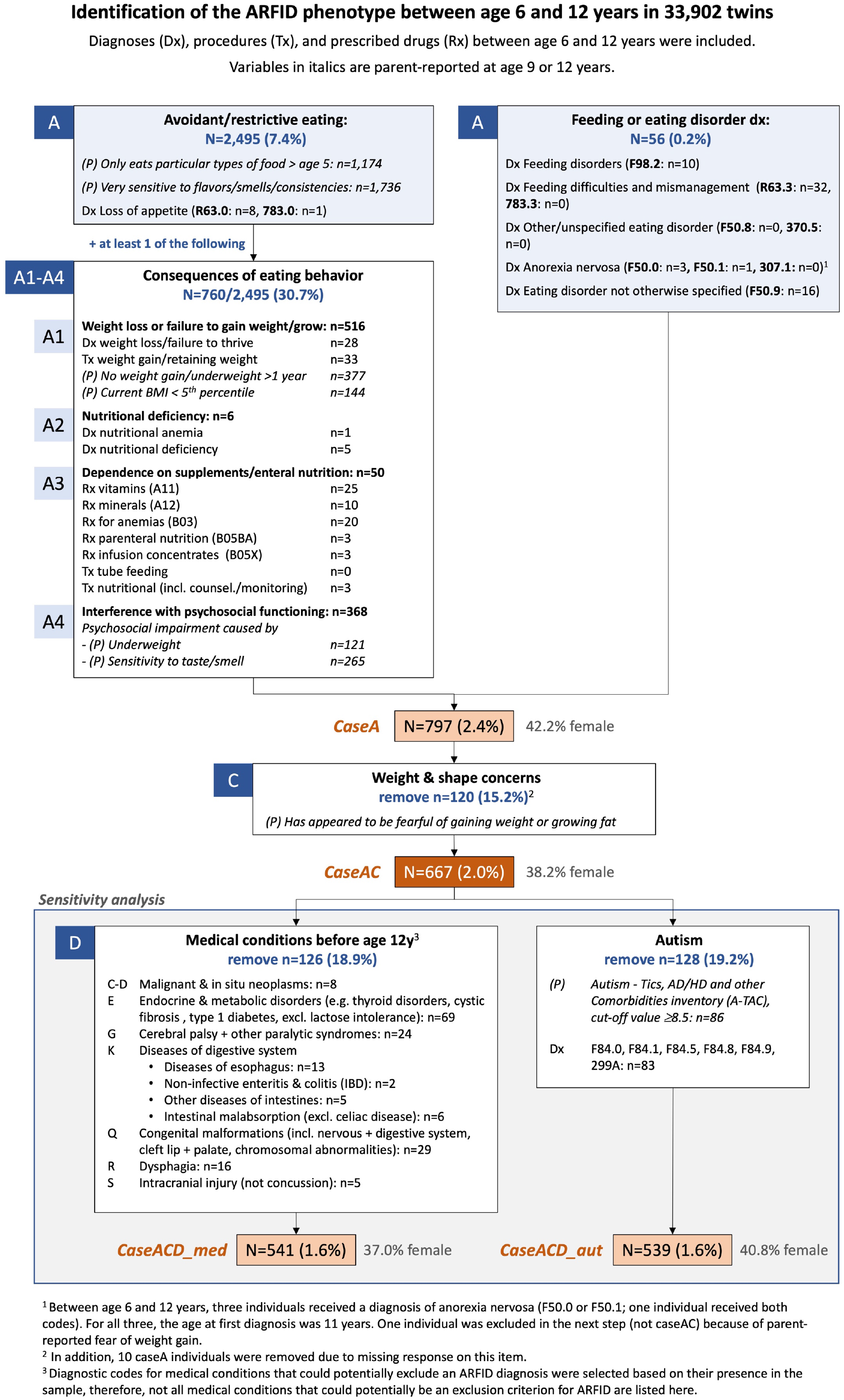
Identification of the ARFID phenotype between age 6 and 12 years in 33,902 twins. Diagnoses (Dx), procedures (Tx), and prescribed drugs (Rx) were included if registered in the National Patient Register or the National Drug Register between age 6 and 12 years. Variables in italics and denoted with *(P)* are based on parent-reports in the Child and Adolescent Twin Study at twin age 9 or 12 years.

All information from CATSS used in this study was reported by parents, either at twin age 9 (87.3% of the sample) or at twin age 12 (12.7% of the sample). To match this age range, we included diagnostic and procedure codes from the NPR, and prescribed drugs from the PDR, to assess *DSM-5 ARFID criterion A* between age 6 and 12 years (we chose age 6 to increase sensitivity for including potential consequences of the eating disturbance diagnosed earlier than age 9, which is the lower age bound for the parent-reports from CATSS). *DSM-5 ARFID criterion C* (eating disturbance not attributable to anorexia nervosa, bulimia nervosa, or body image disturbance) was evaluated using parent-reported fear of weight gain at age 9 or 12. To assess *DSM-5 ARFID criterion D* (eating disturbance not attributable to a concurrent medical condition or another mental disorder), we selected a range of medical conditions <u>at any time before age 12</u> that could potentially explain an eating disturbance. However, in this epidemiological context, it is impossible to determine whether, in each specific case, the selected medical conditions are cause, comorbidity, or consequence of ARFID. Therefore, we did not simply exclude children with the selected medical conditions from the analyses, but rather conducted sensitivity analyses excluding these children. Furthermore, we wanted to ascertain that genetic and environmental influences on the ARFID phenotype are not only due to autism, which is highly heritable^25^ and often co-occurs with ARFID.^26,27^ Therefore, we also conducted sensitivity analyses excluding children with an NPR diagnosis of autism at any point in their life (ICD-9: 299A, ICD-10: F84.0, F84.1, F84.5, F84.8, F84.9), as well as children scoring above cut-off (≥8.5) on the autism scale of the Autism-Tics, AD/HD and other Comorbidities inventory (A-TAC), which has been well-validated for autism.^28-30^ **eTable1** provides a full list of CATSS items, NPR diagnostic and procedure codes, and PDR ATC codes used to evaluate *DSM-5 ARFID criteria A, C, and D*.

In summary, we conducted twin analyses for **four different “case” definitions (Figure 1)**: **(1)** children who only meet *DSM-5 ARFID criterion A* (avoidant/restrictive eating with clinically significant impact that could be due to fear of weight gain in some children, ***caseA***), **(2)** children who meet *DSM-5 ARFID criteria A and C* (excluding children with fear of weight gain, ***caseAC***), **(3)** children who meet *DSM-5 ARFID criteria A, C*, and partially *D* (excluding children with comorbid medical conditions, ***caseACD_med***), and **(4)** children who meet both *DSM-5 ARFID criteria A, C*, and partially *D* (excluding children with comorbid autism, ***caseACD*_*aut***). Since medical conditions and autism are common comorbidities of ARFID,^26,27,31^ we deemed *caseACD_med* and *caseACD_aut* too conversative definitions of ARFID and thus included these in sensitivity analyses. On the other hand, the *caseA* definition is too broad as it does not exclude children with fear of weight gain. We therefore consider *caseAC* to best reflect children with the ARFID phenotype.

### Statistical Analyses

The twin design is based on comparing the relative similarity of MZ and DZ twins on a trait, capitalizing on the fact that MZ twins are genetically identical, whereas DZ twins share on average 50% of their segregating DNA. In contrast to non-twin siblings, twins are also matched for shared environmental influences by sharing the intrauterine environment and growing up in the same family at the same time. By comparing twin correlations, we can therefore estimate the contributions of additive genetics (*A*), shared environment (*C*) or dominant genetic effects (*D*), and non-shared environment (*E*) to a phenotype (albeit *C* and *D* cannot be estimated simultaneously as they confound each other in the classical twin design).

Here, we fitted univariate liability threshold models (which are based on dichotomous data but assume an underlying continuous distribution of liability to the categorical construct) to estimate the relative contribution of genetic and environmental variation to the liability to the ARFID phenotype for each of the four case definitions. As little is known about sex differences in ARFID (including sex differences in its clinical presentation, epidemiology, and etiology), we initially fitted a saturated model including quantitative and qualitative sex limitation to the observed data for all four case definitions (quantitative sex limitation: genetic and environmental variation influences phenotypic variance to differing degrees in females and males; qualitative sex limitation: different genetic and environmental influences in females and males).

Assumption testing for this saturated model revealed no violations of the assumed equal thresholds *across twin order* and *across zygosity* in same-sex twin pairs (**eTable 2;** see number of cases by sex and zygosity in **Table 1**). Twin correlations were estimated from a constrained saturated model, in which the thresholds were equated across twin order and across zygosity within sex (i.e., two thresholds were estimated, one for all females and one for all males). All DZ-ss twin correlations were less than half of the MZ twin correlations, indicating either *D* or sibling contrast effects (−*s;* i.e., parental emphasis on within-pair differences; **Table 2**). Twin correlations of MZ males were somewhat *higher* than twin correlations of MZ females, while twin correlations of DZ-ss males were somewhat *lower* than twin correlations of DZ females, suggesting *quantitative* sex differences. *Qualitative* sex differences were only indicated for one of the four case definitions (*caseACD_med*), where the twin correlation of DZ-os pairs was lower than the average of the twin correlations of DZ-ss females and DZ-ss males. Qualitative sex differences and sibling contrast effects cannot be estimated in the same model as the model would be under-identified. Since there was little indication of *qualitative* sex differences, we fitted ADE-s models with only *quantitative* sex limitation. Sibling contrast effects were modeled by adding a causal pathway (*-s*) between one twin’s phenotype and their co-twin’s phenotype. Significance of individual parameters was tested by constraining them to be equal to zero. The best-fitting models were chosen based on the likelihood ratio test (the reduced model was favored if model fit did not deteriorate significantly). Data management was performed using SAS 9.4 software. Data analysis was performed in and OpenMx version 2.20.6^32^ in R 4.2.0 software.

**Table 1.** Number of cases with the ARFID phenotype and proportion of cases meeting sub-criteria of DSM-5 ARFID criterion A by sex, zygosity, and case definition, n (%)

|  | **caseA** | **caseAC** | **caseACD_med** | **caseACD_aut** |
| --- | --- | --- | --- | --- |
| **Number of cases** |  |  |  |  |
| Total sample (N = 33,902) | 797 (2.4%) | **667 (2.0%)** | 541 (1.6%) | 539 (1.6%) |
| By sex |  |  |  |  |
| Males (N = 17,151) | 461 (2.7%) | **412 (2.4%)** | 341 (2.0%) | 319 (1.9%) |
| Females (N = 16,751) | 336 (2.0%) | **255 (1.5%)** | 200 (1.2%) | 220 (1.3%) |
| By sex and zygosity |  |  |  |  |
| MZ males (N = 4,984) | 116 (2.3%) | **97 (2.0%)** | 79 (1.6%) | 75 (1.5%) |
| MZ females (N = 5,384) | 95 (1.8%) | **75 (1.4%)** | 51 (0.9%) | 65 (1.2%) |
| DZ-ss males (N = 6,336) | 166 (2.6%) | **149 (2.4%)** | 125 (2.0%) | 111 (1.8%) |
| DZ-ss females (N = 5,536) | 125 (2.3%) | **103 (1.9%)** | 87 (1.6%) | 88 (1.6%) |
| DZ-os males (N = 5,831) | 179 (3.1%) | **166 (2.9%)** | 137 (2.4%) | 133 (2.3%) |
| DZ-os females (N = 5,831) | 116 (2.0%) | **77 (1.3%)** | 62 (1.1%) | 67 (1.2%) |
| **Proportion of cases meeting sub-criteria of DSM-5 ARFID criterion A** |  |  |  |  |
| A1 - low weight/failure to thrive | 535 (67.1%) | **445 (66.7%)** | 349 (64.5%) | 345 (64.0%) |
| A2 - nutritional deficiency | 6 (0.8%) | **4 (0.6%)** | 2 (0.4%) | 2 (0.4%) |
| A3 - nutritional supplements | 54 (6.8%) | **49 (7.3%)** | 29 (5.4%) | 30 (5.6%) |
| A4 - psychosocial impairment | 375 (47.2%) | **341 (51.4%)** | 291 (54.1%) | 279 (52.1%) |

caseA: children who only meet DSM-5 ARFID criterion A (avoidant restrictive eating with clinically significant impact that could be due to fear of weight gain in some children; caseAC: children who meet DSM-5 ARFID criteria A and C (excluding children with fear of weight gain; caseACD\_med: children who meet DSM-5 ARFID criteria A, C, and partially D (excluding children with comorbid medical conditions); caseACD\_aut: children who meet both DSM-5 ARFID criteria A, C, and partially D (excluding children with comorbid autism).

**Table 2.** Tetrachoric twin correlations and proband-wise concordance rates for the ARFID phenotype by zygosity, sex, and case definition

|  | Male |  | Female |  | DZ-os |
| --- | --- | --- | --- | --- | --- |
|  | MZ | DZ-ss | MZ | DZ-ss |  |
| Number of pairs | 2,492 | 3,168 | 2,692 | 2,768 | 5,831 |
| Twin correlations, *r* (95% CI) |  |  |  |  |  |
| caseA | 0.77 (0.66-0.85) | 0.18 (-0.03-0.37) | 0.63 (0.46-0.76) | 0.31 (0.11-0.49) | 0.24 (0.10-0.37) |
| caseAC | 0.76 (0.64-0.85) | 0.17 (-0.05-0.37) | 0.69 (0.52-0.81) | 0.28 (0.06-0.48) | 0.23 (0.06-0.38) |
| caseACD_med | 0.78 (0.65-0.87) | 0.24 (0.01-0.44) | 0.59 (0.33-0.77) | 0.28 (0.03-0.50) | 0.12 (-0.10-0.32) |
| caseACD_aut | 0.73 (0.57-0.84) | 0.13 (-0.15-0.38) | 0.67 (0.49-0.81) | 0.19 (-0.10-0.43) | 0.17 (-0.03-0.35) |
| Proband-wise concordance rates |  |  |  |  |  |
| caseA | 0.38 | 0.06 | 0.23 | 0.10 | 0.07 |
| caseAC | 0.35 | 0.05 | 0.27 | 0.08 | 0.06 |
| caseACD_med | 0.35 | 0.06 | 0.16 | 0.07 | 0.03 |
| caseACD_aut | 0.29 | 0.04 | 0.25 | 0.05 | 0.04 |

Probandwise concordance rates were calculated as 2 x number of concordant pairs / ([2 x number of concordant pairs] + number of discordant pairs). They indicate the probability that a co-twin of a proband is also a proband.
caseA: children who only meet DSM-5 ARFID criterion A (avoidant restrictive eating with clinically significant impact that could be due to fear of weight gain in some children; caseAC: children who meet DSM-5 ARFID criteria A and C (excluding children with fear of weight gain; caseACD\_med: children who meet DSM-5 ARFID criteria A, C, and partially D (excluding children with comorbid medical conditions); caseACD\_aut: children who meet both DSM-5 ARFID criteria A, C, and partially D (excluding children with comorbid autism)

## Results

### Identification of the ARFID phenotype

In our sample of 33,902 individuals, we identified 797 children (2.4%) who had the ARFID *caseA* phenotype (i.e., not considering potential fear of weight gain) (**Figure 1, Table 1**). After excluding children with parent-reported fear of weight gain, 667 children were classified as having the ARFID *caseAC* phenotype, corresponding to a population prevalence of 2.0% (38.2% female). Two thirds (445/667, 66.7%) of children with the ARFID *caseAC* phenotype met *DSM-5 ARFID criterion A1* (low weight/failure to gain thrive), and 51.4% (341/667) met *DSM-5 ARFID criterion A4* (psychosocial impairment; **Table 1**). Only a small minority met *DSM-5 ARFID criterion A2* (nutritional deficiency, 4/667, 0.6%) or *DSM-5 ARFID criterion A3* (nutritional supplements, 49/667, 7.3%). For the sensitivity analyses, we further excluded 126 children with medical conditions that could potentially explain the eating disturbance (*caseACD*, n=541, 1.6%) and 128 children with autism (*caseACD_aut*, n=539, 1.6%).

### Model fitting and heritability of the ARFID phenotype

According to likelihood ratio tests, model fits did not deteriorate significantly when quantitative sex limitation was dropped (**Table 3**). In addition, the ADE-s models *including* quantitative sex limitation were severely underpowered, as indicated by the large confidence intervals (CIs) for the A and D components, which also included zero for all case definitions (**Table 4**). We therefore fitted nested models of ADE-s models *without* sex limitation (**eTable 3**). AE-s models showed the best fit for all four case definitions (**Table 3**). Heritability of the ARFID *caseAC* phenotype was 0.79 (95% CI 0.71-0.86); with small but statistically significant non-shared environmental effects (0.21, 95% CI 0.14-0.29) and sibling contrast effects (−0.09, 95% CI -0.14 - -0.04; **Table 4**). Heritability was very similar across all four case definitions (point estimates: 0.77-0.80).

**Table 3.**
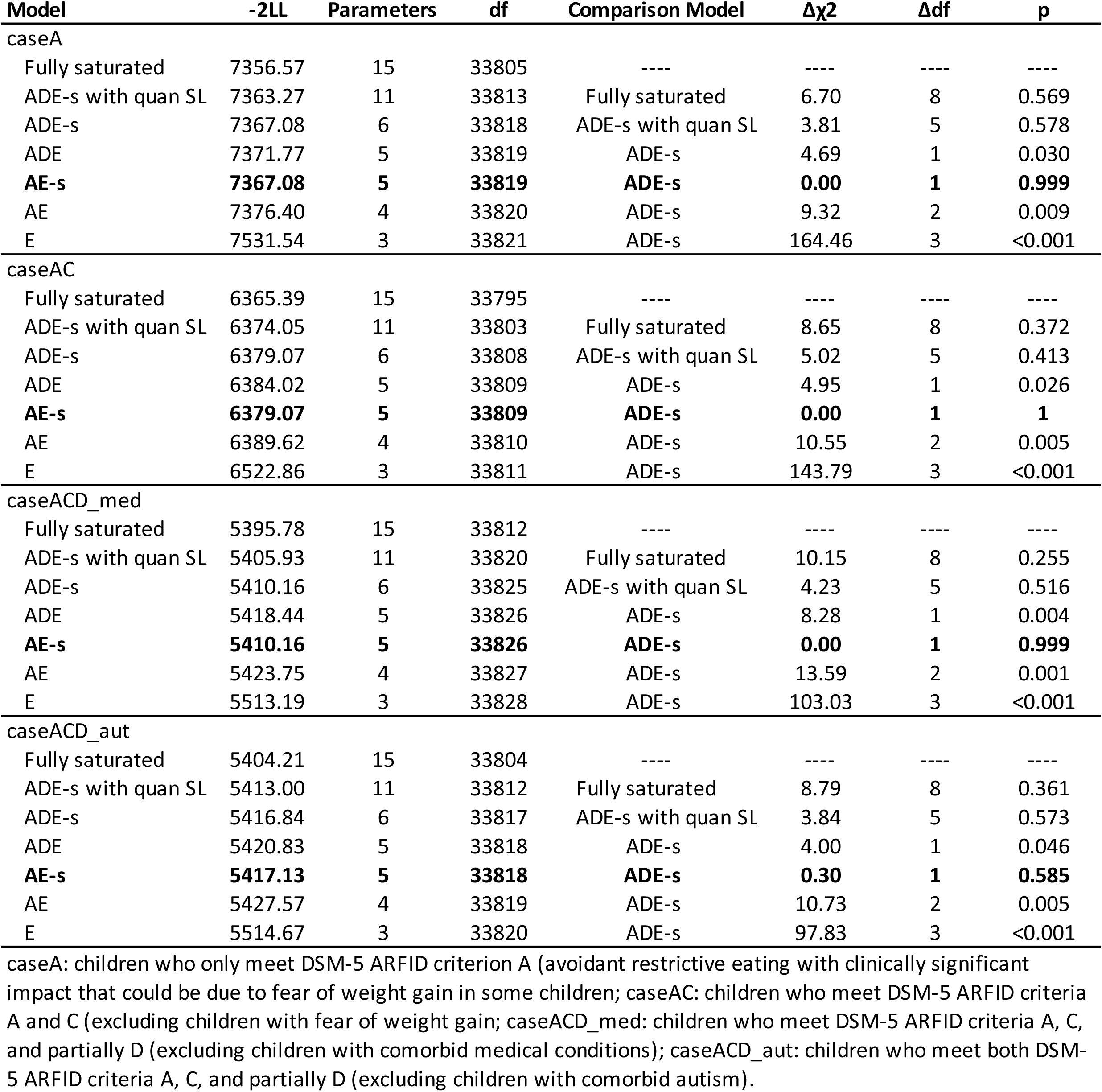
Model fit statistics for ADE-s models with qualitative and quantitative sex limitation (quan SL) and nested models by case definition

| Model | -2LL | Parameters | df | Comparison Model | $\Delta\chi^2$ | $\Delta df$ | p |
| --- | --- | --- | --- | --- | --- | --- | --- |
| caseA |  |  |  |  |  |  |  |
| Fully saturated | 7356.57 | 15 | 33805 | ---- | ---- | ---- | ---- |
| ADE-s with quan SL | 7363.27 | 11 | 33813 | Fully saturated | 6.70 | 8 | 0.569 |
| ADE-s | 7367.08 | 6 | 33818 | ADE-s with quan SL | 3.81 | 5 | 0.578 |
| ADE | 7371.77 | 5 | 33819 | ADE-s | 4.69 | 1 | 0.030 |
| **AE-s** | **7367.08** | **5** | **33819** | **ADE-s** | **0.00** | **1** | **0.999** |
| AE | 7376.40 | 4 | 33820 | ADE-s | 9.32 | 2 | 0.009 |
| E | 7531.54 | 3 | 33821 | ADE-s | 164.46 | 3 | <0.001 |
| caseAC |  |  |  |  |  |  |  |
| Fully saturated | 6365.39 | 15 | 33795 | ---- | ---- | ---- | ---- |
| ADE-s with quan SL | 6374.05 | 11 | 33803 | Fully saturated | 8.65 | 8 | 0.372 |
| ADE-s | 6379.07 | 6 | 33808 | ADE-s with quan SL | 5.02 | 5 | 0.413 |
| ADE | 6384.02 | 5 | 33809 | ADE-s | 4.95 | 1 | 0.026 |
| **AE-s** | **6379.07** | **5** | **33809** | **ADE-s** | **0.00** | **1** | **1** |
| AE | 6389.62 | 4 | 33810 | ADE-s | 10.55 | 2 | 0.005 |
| E | 6522.86 | 3 | 33811 | ADE-s | 143.79 | 3 | <0.001 |
| caseACD_med |  |  |  |  |  |  |  |
| Fully saturated | 5395.78 | 15 | 33812 | ---- | ---- | ---- | ---- |
| ADE-s with quan SL | 5405.93 | 11 | 33820 | Fully saturated | 10.15 | 8 | 0.255 |
| ADE-s | 5410.16 | 6 | 33825 | ADE-s with quan SL | 4.23 | 5 | 0.516 |
| ADE | 5418.44 | 5 | 33826 | ADE-s | 8.28 | 1 | 0.004 |
| **AE-s** | **5410.16** | **5** | **33826** | **ADE-s** | **0.00** | **1** | **0.999** |
| AE | 5423.75 | 4 | 33827 | ADE-s | 13.59 | 2 | 0.001 |
| E | 5513.19 | 3 | 33828 | ADE-s | 103.03 | 3 | <0.001 |
| caseACD_aut |  |  |  |  |  |  |  |
| Fully saturated | 5404.21 | 15 | 33804 | ---- | ---- | ---- | ---- |
| ADE-s with quan SL | 5413.00 | 11 | 33812 | Fully saturated | 8.79 | 8 | 0.361 |
| ADE-s | 5416.84 | 6 | 33817 | ADE-s with quan SL | 3.84 | 5 | 0.573 |
| ADE | 5420.83 | 5 | 33818 | ADE-s | 4.00 | 1 | 0.046 |
| **AE-s** | **5417.13** | **5** | **33818** | **ADE-s** | **0.30** | **1** | **0.585** |
| AE | 5427.57 | 4 | 33819 | ADE-s | 10.73 | 2 | 0.005 |
| E | 5514.67 | 3 | 33820 | ADE-s | 97.83 | 3 | <0.001 |

caseA: children who only meet DSM-5 ARFID criterion A (avoidant restrictive eating with clinically significant impact that could be due to fear of weight gain in some children; caseAC: children who meet DSM-5 ARFID criteria A and C (excluding children with fear of weight gain; caseACD\_med: children who meet DSM-5 ARFID criteria A, C, and partially D (excluding children with comorbid medical conditions); caseACD\_aut: children who meet both DSM-5 ARFID criteria A, C, and partially D (excluding children with comorbid autism).

**Table 4.** Variance component estimates for (A) ADE-s models with quantitative sex limitation and (B) the final model (AE-s without sex limitation)

|  | Males |  |  | Females |  |  | Sibling contrast effects (-s) |  |  |
| --- | --- | --- | --- | --- | --- | --- | --- | --- | --- |
|  | A | D | E | A | D | E | Same sex males | Same sex females | Opposite sex twins |
| caseA | 0.70 (0.00-0.89) | 0.13 (0.00-0.85) | 0.18 (0.11-0.29) | 0.65 (0.00-0.81) | 0.03 (0.00-0.68) | 0.32 (0.19-0.60) | -0.10 (-0.17-0.01) | -0.04 (-0.13-0.09) | -0.04 (-0.15-0.31) |
| caseAC | 0.83 (0.00-0.90) | 0.00 (0.00-0.83) | 0.17 (0.10-0.26) | 0.56 (0.00-0.83) | 0.14 (0.00-0.76) | 0.30 (0.17-0.55) | -0.13 (-0.19--0.04) | -0.01 (-0.11-0.10) | -0.06 (-0.17-0.29) |
| caseACD_med | 0.85 (0.32-0.91) | 0.00 (0.00-0.00) | 0.15 (0.09-0.25) | 0.67 (0.02-0.83) | 0.02 (0.00-0.74) | 0.31 (0.17-0.62) | -0.12 (-0.18--0.04) | -0.08 (-0.19-0.06) | -0.13 (-0.23-0.28) |
| caseACD_aut | 0.80 (0.00-0.88) | 0.01 (0.00-0.83) | 0.20 (0.12-0.32) | 0.28 (0.00-0.81) | 0.41 (0.00-0.80) | 0.31 (0.18-0.58) | -0.12 (-0.19--0.03) | -0.02 (-0.13-0.10) | -0.04 (-0.20-0.29) |

*(B) AE-s without sex limitation (final model)*

|  | A | D | E | s |
| --- | --- | --- | --- | --- |
| caseA | 0.78 (0.69-0.84) | ----- | 0.22 (0.16-0.31) | -0.09 (-0.13- -0.03) |
| caseAC | 0.79 (0.71-0.86) | ----- | 0.21 (0.14-0.29) | -0.09 (-0.14- -0.04) |
| caseACD_med | 0.80 (0.71-0.86) | ----- | 0.20 (0.14-0.29) | -0.12 (-0.16- -0.06) |
| caseACD_aut | 0.77 (0.67-0.84) | ----- | 0.23 (0.16-0.33) | -0.10 (-0.16- -0.05) |

caseA: children who only meet DSM-5 ARFID criterion A (avoidant restrictive eating with clinically significant impact that could be due to fear of weight gain in some children; caseAC: children who meet DSM-5 ARFID criteria A and C (excluding children with fear of weight gain; caseACD\_med: children who meet DSM-5 ARFID criteria A, C, and partially D (excluding children with comorbid medical conditions); caseACD\_aut: children who meet both DSM-5 ARFID criteria A, C, and partially D (excluding children with comorbid autism).

## Discussion

In light of the lack of large-scale epidemiological twin data on ARFID, we leveraged existing data to create four definitions of an ARFID phenotype. Combining data from parent-reports and national health registers, we identified 667 children (2%) with the ARFID phenotype and found that the ARFID phenotype is highly heritable. ARFID heritability was 0.79 (95% CI 0.71-0.86), placing it amongst the most heritable of psychiatric disorders (e.g., autism 0.64-0.91^33^; schizophrenia 0.79^34^; attention deficit hyperactivity disorder [ADHD] 0.77-0.88;^35^ and bipolar disorder 0.50-0.71^36^). Moreover, the heritability of the ARFID phenotype was higher than that of other eating disorders, namely anorexia nervosa (0.48-0.74), bulimia nervosa (0.55-0.61), and binge-eating disorder (0.39-0.45).^21^ Our results extend and confirm previous twin studies of other feeding-related phenotypes of moderate to high heritability such as appetite (.53-.84),^19^ food fussiness (.46-.78),^14,16^ an food neophobia (.58-.78).^16-18^ Sensitivity analyses excluding individuals with autism and medical conditions that could potentially explain the eating disturbance led to only very minor changes in heritability estimates, suggesting that these conditions did not account for the high heritability. Interestingly, our twin models revealed sibling contrast effects for ARFID, which are commonly observed in neurodevelopmental disorders including autism and ADHD,^37,38^ suggesting that parents’ ratings of their twins’ eating problems might amplify differences between their twins. Modeling these contrast effects led to an increase in the heritability estimate from 0.68 (in the AE model) to 0.79 (in the AE-s model). Qualitative sex differences (i.e., *different* genetic and environmental influences in males vs. females) did not seem to play an important role, whereas there was some indication for a *higher* heritability in males (i.e., quantitative sex difference). Although these were not significant, our models including sex limitations were underpowered and sex differences therefore need to be tested in future studies with larger sample sizes.

To construct the ARFID phenotype we were limited to existing data in CATSS, the NPR, and the PDR. Most cases (760/797, 95%) were identified via the parent-reported gate items “only wanted to eat particular types of food during a period after age 5” and “particularly sensitive to certain flavors, smells, or consistencies” (as opposed to being identified with a feeding or eating disorder between age 6 and 12; **Figure 1**). Therefore, the ARFID phenotype derived in this study is likely to reflect cases that include a *sensory-based avoidance* component (typically associated with selective eating). This is relevant as genetic and environmental influences might be differentially implicated across predominant ARFID presentations, for instance, the ARFID phenotype in people who had adverse conditioning experiences such as choking on food might have a larger environmental contribution. Sensory-based avoidance is the most common presentation (62-73%) of ARFID in children,^8,39^ and the presentations are in no way mutually exclusive. More than half of children with ARFID have mixed presentations of sensory-based avoidance with fear-based avoidance or sensory-based avoidance with lack of interest.^39^ Indeed, a high proportion of ARFID identified in this study was via low weight and failure to thrive (535/797, 67%), which are more commonly associated with lack of interest and fear-based avoidance.^40,41^ Future studies aimed at delineating differences in biological and environmental risk factors based on predominant clinical characteristics will require larger samples and more extensive phenotyping.

### Strengths and Limitations

Our study had several strengths. We optimized existing data resources to provide the first heritability estimates of ARFID based on a sample size larger than typically reported in single site clinical samples. By triangulating questionnaire and health register data we accessed many different indicators of ARFID to carefully define the phenotype and specify exclusions for sensitivity analyses. Although our algorithm-derived definition of ARFID has not been validated, the prevalence and sex distribution were consistent with available published estimates (prevalence 0.3-3.2%,^8,42,43^ sex distribution ∼1:1, with some studies finding a slight female preponderance^8,42^ and others finding a slight male preponderance^44,45^), providing some confidence in the phenotype. Our study focused on ARFID in children aged 6-12 years, yet the disorder is not confined to the childhood years.^46,47^ Subsequent studies using different designs and samples should also include adults to further characterize this illness across the lifespan. Finally, even with a sample of ∼34,000 twins, analyses were underpowered for testing sex differences. Future research should estimate ARFID heritability in even larger samples including older individuals by using validated measures appropriate for epidemiological studies, which are expected to be available in the upcoming years.

## Conclusions

This study shows that, given the similar prevalence figures and sex distribution, existing register-based epidemiological data may be used to approximate ARFID, and that the resulting broad ARFID phenotype is highly heritable—with significant contributions from non-shared environmental factors—and distinguishable from other eating disorders characterized by fear of weight gain and older average age of onset. The high heritability of the ARFID phenotype provides strong support for future twin and molecular genetic studies of ARFID.

## Data Availability

Due to ethical restrictions related to protecting confidentiality connected to the Swedish Twin Registry, relevant data are available on request pending approval from the Swedish Twin Registry.

## Acknowledgements

We gratefully acknowledge the contribution of the participants in the Child and Adolescent Twin Study in Sweden (CATSS) and their families. We acknowledge the Swedish Twin Registry for access to data. The Swedish Twin Registry is managed by Karolinska Institutet and receives funding through the Swedish Research Council under the grant no 2017-00641.

## Author Contributions

Dr Dinkler had full access to all of the data in the study and takes responsibility for the integrity of the data and the accuracy of the data analysis.

### Concept and design

Dinkler, Taylor, Bulik.

### Acquisition, analysis, or interpretation of data

All authors.

### Drafting of the manuscript

Dinkler.

### Critical revision of the manuscript for important intellectual content

All authors.

### Statistical analysis

Dinkler, Taylor.

### Obtained funding

Dinkler, Bulik, Lichtenstein, Lundström, Larsson.

### Administrative, technical, or material support

Lichtenstein, Lundström, Larsson.

### Supervision

Taylor, Bulik.

## Supplementary material

**eTable 1**. Variables from the Child and Adolescent Twin Study in Sweden (parent-reports), the National Patient Register (diagnostic and procedure codes), and the Prescribed Drug Register (ATC codes) used to evaluate DSM-5 ARFID criteria A, C, and D

**eTable 2**. Assumption testing for models with quantitative sex limitation by case definition

**eTable 3**. Variance component estimates for ADE-s models <u>without sex limitation</u> and nested models by case definition (final models in bold)

