## Supplemental Tables 1-3 for "Genetic and environmental etiology of the broad avoidant restrictive food intake disorder phenotype in 6- to-12-year-old Swedish twins"

**eTable 1**. Variables from the Child and Adolescent Twin Study in Sweden (parent-reports), the National Patient Register (diagnostic and procedure codes), and the Prescribed Drug Register (ATC codes) used to evaluate DSM-5 ARFID criteria A, C, and D

| DSM-5 ARFID criterion | Name in Figure 1 | Description of diagnosis, procedure, prescribed medication, or parent-reports | Included codes for diagnosis (ICD-9/ICD-10), procedure, prescribed medication (ATC); and response options for parent-reports | Exclusions |
|---|---|---|---|---|
| A - Feeding or eating disorder dx | Dx Feeding disorders | Feeding disorder of infancy or childhood | F98.2 | ----- |
|  | Dx Feeding difficulties and mismanagement | Feeding difficulties and mismanagement | 783.3, R63.3 | ----- |
|  | Dx Other/unspecified eating disorder | Other and unspecified disorders of eating | 307.5, F50.8 | ----- |
|  | Dx Anorexia nervosa | Anorexia nervosa | 307.1, F50.0, F50.1 | ----- |
|  | Dx Eating disorder not otherwise specified | Eating disorder not otherwise specified (EDNOS) | F50.9 | ----- |
| A - Avoidant/ restrictive eating | (P) Only eats particular types of food > age 5 | Has he/she ever had a period after age 5 when he/she only wanted to eat particular types of food? | Yes<br>Yes, to some extent | No |
| | (P) Very sensitive to flavors/smells/consistencies | Is he/she particularly sensitive to certain flavours, smells, or consistencies? | Yes | Yes, to some extent<br>No |
| | Dx Loss of appetite | Anorexia (Loss of appetite) | 783.0, R63.0 | |
| A1 | Dx weight loss/failure to thrive | Other lack of expected normal physiological development in childhood (failure to gain weight/thrive) | 783.4, R62.8 | ----- |
|  |  | Abnormal weight loss | 783.2, R63.4 | ----- |
|  | Tx weight gain/retaining weight | Weight measurement | AV112 | ----- |
|  |  | Assessment of weight maintenance functions | PE006 | ----- |
|  |  | Support for weight gain | QE010 | ----- |
| | (P) No weight gain/underweight >1 year | Has he/she ever failed to gain enough weight for more than a year or been underweight? | Yes<br>Yes, to some extent | No |
| | (P) Current BMI < -5th percentile | BMI calculated as kg/m2 based on parent-reported height and weight of the twin; BMI < 5th percentile defined by sex and age based on the current sample (male age 9: n=13,858; female age 9: 13,634; male age 12: n=3,293; female age [illegible] | BMI < -2SD | |
| A2 | Dx nutritional anemia | Nutritional anemias | 280.1, 280.8, 280.9, 281<br>D50.1, D50.8, D50.9, D51.3, D51.8, D51.9, D52.0, D52.8, D52.9, D53 | D50.0, D51.0, D51.1, D52.2, D52.1 (deficiencies due to blood loss, malabsorption, and intrinsic factors; drug-induced deficiencies) |
| | Dx nutritional deficiency | Malnutrition & other nutritional deficiencies | 260-269, E40-E46, E50-E64 | 268, E55 (Vitamin D deficiency) |
| A3 | Rx vitamins | Prescribed vitamins | A11 | A11CC (Vitamin D) |
| | Rx minerals | Prescribed minerals | A12 | |
| | Rx for anemias | Prescriptions for anemias | B03 | |
| | Rx parenteral nutrition | Prescribed parenteral nutrition | B05BA | |
| | Rx infusion concentrates | Prescribed infusion concentrates | B05X | |
| | Tx tube feeding | Enteral nutrition treatment via tube | DJ010 | |
| | | Tube feeding | DV065 | |
| | | Nasogastric or nasogastroduodenal tube | TJD00 | |
| | | Other tube in ventricle or duodenum | TJD10 | |
| | | Changing the gastrostomy catheter | TJD20 | |
| | | Jejunal tube via gastrostomy | TJF10 | |
| | | Attention to gastrostomy | V55.1, Z43.1 | |
| | | Gastrostomy status | V44.1, Z93.1 | |
| | Tx nutritional (incl. counsel./monitoring) | Nutritional value calculation | AV090-AV092 | |
| | | Eating training | DJ012 | |
| | | Dietary supplements, dietician assessment | DV043 | |
| | | Nutritional treatment | DV051-DV056 | |
| | | Monitoring of nutritional intake | QE003 | |

| | | | | |
|---|---|---|---|---|
| | | Feeding | QN022 | |
| | | Dietician | XS912 | |
| | | Dietary counselling and surveillance | Z71.3 | |
| A4 | Psychosocial impairment caused by (P) Underweight | Have peculiarities or problems relating to underweight caused significant impairment in school, among peers or at home? | Yes<br>Yes, to some extent | No |
| | | Do the peculiarities or problems relating to underweight cause him/her significant suffering? | Yes<br>Yes, to some extent | No |
| | Psychosocial impairment caused by (P) Sensitivity to taste/smell | Have the problems relating to sensitivity to flavours, smells, or consistencies caused significant impairment in school, among peers or at home? | Yes<br>Yes, to some extent | No |
| | | Do the problems relating to sensitivity to flavours, smells, or consistencies cause him/her significant suffering? | Yes<br>Yes, to some extent | No |
| C - Weight & shape concerns | Weight & shape concerns | Has he/she appeared to be fearful of gaining weight or becoming fat? | Yes<br>Yes, to some extent | No |
| D - Medical conditions before age 12y[1] | Malignant & in situ neoplasms | Malignant neoplasms | 140–209, C00–C97 | |
| | | In situ neoplasms | 230–234, D00–D09 | |
| | Endocrine & metabolic disorders | Disorders of thyroid gland | 240-246, E00-E07 | |
| | | Diabetes mellitus | 249-250, E10-E14 | |
| | | Other disorders of glucose regulation and pancreatic internal secretion | 251, E15-E16 | |
| | | Disorders of other endocrine glands | 249-259, E20-E35 | |
| | | Metabolic disorders | 270-279, E70-E90 | 271.3, E73 (lactose intolerance) |
| | Cerebral palsy + other paralytic syndromes | Cerebral palsy and other paralytic syndromes | 342-344, G80–G83 | |
| | Diseases of digestive system - Diseases of esophagus | Oesophagitis | 530, K20 | |
| | | Gastro-oesophageal reflux disease with oesophagitis | 530, K21.0 | |
| | | Other diseases of oesophagus | 530, K22 | |
| | Diseases of digestive system - Non-infective enteritis & colitis (IBD) | Crohn disease [regional enteritis] | 555, K50 | |
| | | Ulcerative colitis | 556, K51 | |
| | Diseases of digestive system - Other diseases of intestines | Paralytic ileus and intestinal obstruction without hernia | 560, K56 | |
| | | Neurogenic bowel, not elsewhere classified | 564.8, K59.2 | |
| | | Perforation of intestine (nontraumatic) | 569.4, 569.8, K63.1 | |
| | Diseases of digestive system - Intestinal malabsorption | Intestinal malabsorption | 579, K90 | 579.0, K90.0 (celiac disease) |
| | Birth injury to central nervous system | Intracranial laceration and haemorrhage due to birth injury | 767.0, P10 | |
| | | Other birth injuries to central nervous system | 767.4, 767.5, 767.7, P11 | |
| | Congenital malformations | Congenital malformations of the nervous system | 740-742, Q00-Q07 | |
| | | Cleft lip and cleft palate | 749, Q35-Q37 | |
| | | Other congenital malformations of the digestive system | 750-751, Q38-Q45 | |
| | | Chromosomal abnormalities, not elsewhere classified | 758-759, Q90-Q99 | |
| | Dysphagia | Dysphagia | 787.2, R13 | |
| | Intracranial injury | Intracranial injury | 850.0-854.1, S06.0–S06.9 | 850, S06.0 (concussion) |
| D - Autism | A-TAC | Autism - Tics, AD/HD and other Comorbidities inventory (A-TAC), 17 items for autism | Theoretical range: 0-17, cut-off value: >=8.5 | |
| | Dx | Pervasive developmental disorders | 299A, F84.0, F84.1, F84.5, F84.8, F84.9 | |

[1] Diagnostic codes for medical conditions that could potentially exclude an ARFID diagnosis were selected based on their presence in the sample, therefore, not *all* medical conditions that could potentially be an exclusion criterion for ARFID are listed here.

**eTable 2**. Assumption testing for models with qualitative and quantitative sex limitation by case definition

| Model | -2LL | Parameters | df | Δχ2 | Δdf | p |
|---|---|---|---|---|---|---|
| caseA | | | | | | |
| Fully Saturated | 7356.6 | 15 | 33805 | ---- | ---- | ---- |
| Equal thresholds within same-sex twin pairs | 7360.10 | 11 | 33809 | 3.53 | 4 | 0.474 |
| Equal thresholds across zygosity for same-sex twin pair: | 7363.9 | 9 | 33811 | 7.28 | 6 | 0.296 |
| caseAC | | | | | | |
| Fully Saturated | 6365.4 | 15 | 33795 | ---- | ---- | ---- |
| Equal thresholds within same-sex twin pairs | 6368.4 | 11 | 33799 | 3.03 | 4 | 0.552 |
| Equal thresholds across zygosity for same-sex twin pair: | 6373.4 | 9 | 33801 | 8.05 | 6 | 0.235 |
| caseACD_med | | | | | | |
| Fully Saturated | 5395.8 | 15 | 33812 | ---- | ---- | ---- |
| Equal thresholds within same-sex twin pairs | 5398.4 | 11 | 33816 | 2.66 | 4 | 0.617 |
| Equal thresholds across zygosity for same-sex twin pair: | 5408.1 | 9 | 33818 | 12.34 | 6 | 0.055 |
| caseACD_aut | | | | | | |
| Fully Saturated | 5404.2 | 15 | 33804 | ---- | ---- | ---- |
| Equal thresholds within same-sex twin pairs | 5407.2 | 11 | 33808 | 2.98 | 4 | 0.562 |
| Equal thresholds across zygosity for same-sex twin pair: | 5410.7 | 9 | 33810 | 6.43 | 6 | 0.376 |

caseA: children who only meet DSM-5 ARFID criterion A (avoidant restrictive eating with clinically significant impact that could be due to fear of weight gain in some children; caseAC: children who meet DSM-5 ARFID criteria A and C (excluding children with fear of weight gain; caseACD_med: children who meet DSM-5 ARFID criteria A, C, and partially D (excluding children with comorbid medical conditions); caseACD_aut: children who meet both DSM-5 ARFID criteria A, C, and partially D (excluding children with comorbid autism).

**eTable3.** Variance component estimates for ADE-s models without sex limitation and nested models by case definition (final models in bold)

| | Variance component | | | |
|---|---|---|---|---|
| | A | D | E | s |
| caseA | | | | |
| ADE-s | 0.78 (0.15-0.84) | 0.00 (0.00-0.00) | 0.22 (0.16-0.32) | -0.09 (-0.13- -0.01) |
| ADE | 0.25 (0.00-0.65) | 0.46 (0.04-0.78) | 0.29 (0.21-0.38) | ----- |
| **AE-s** | **0.78 (0.69-0.84)** | **-----** | **0.22 (0.16-0.31)** | **-0.09 (-0.13- -0.03)** |
| AE | 0.67 (0.58-0.75) | ----- | 0.33 (0.25-0.42) | ----- |
| caseAC | | | | |
| ADE-s | 0.79 (0.04-0.86) | 0.00 (0.00-0.73) | 0.21 (0.14-0.30) | -0.09 (-0.14- -0.01) |
| ADE | 0.17 (0.00-0.61) | 0.56 (0.09-0.81) | 0.27 (0.19-0.36) | ----- |
| **AE-s** | **0.79 (0.71-0.86)** | **-----** | **0.21 (0.14-0.29)** | **-0.09 (-0.14- -0.04)** |
| AE | 0.69 (0.59-0.77) | ----- | 0.31 (0.23-0.41) | ----- |
| caseACD_med | | | | |
| ADE-s | 0.80 (0.07-0.86) | 0.00 (0.00-0.00) | 0.20 (0.14-0.30) | -0.12 (-0.16- -0.04) |
| ADE | 0.09 (0.00-0.60) | 0.63 (0.09-0.71) | 0.28 (0.19-0.39) | ----- |
| **AE-s** | **0.80 (0.71-0.86)** | **-----** | **0.20 (0.14-0.29)** | **-0.12 (-0.16- -0.06)** |
| AE | 0.66 (0.55-0.76) | ----- | 0.34 (0.24-0.45) | ----- |
| caseACD_aut | | | | |
| ADE-s | 0.51 (0.00-0.83) | 0.25 (0.00-0.81) | 0.24 (0.16-0.35) | -0.08 (-0.15-0.00) |
| ADE | 0.00 (0.00-0.00) | 0.70 (0.17-0.79) | 0.30 (0.21-0.41) | ----- |
| **AE-s** | **0.77 (0.67-0.84)** | **-----** | **0.23 (0.16-0.33)** | **-0.10 (-0.16- -0.05)** |
| AE | 0.64 (0.52-0.74) | ----- | 0.36 (0.26-0.48) | ----- |

caseA: children who only meet DSM-5 ARFID criterion A (avoidant restrictive eating with clinically significant impact that could be due to fear of weight gain in some children; caseAC: children who meet DSM-5 ARFID criteria A and C (excluding children with fear of weight gain; caseACD_med: children who meet DSM-5 ARFID criteria A, C, and partially D (excluding children with comorbid medical conditions); caseACD_aut: children who meet both DSM-5 ARFID criteria A, C, and partially D (excluding children with comorbid autism).
